# Partial breast irradiation after lumpectomy with omission of surgical axillary evaluation

**DOI:** 10.64898/2026.06.29.26356836

**Authors:** Diana A. Roth O’Brien, Lillian A. Boe, Boris A. Mueller, Giacomo Montagna, Emma N. Hahesy, John J. Cuaron, J. Isabelle Choi, Michael B. Bernstein, Beryl McCormick, Simon N. Powell, Atif J. Khan, Lior Z. Braunstein

**Author notes:** **Correspondence:** Lior Z. Braunstein, MD, Memorial Sloan Kettering Cancer Center, 1275 York Ave, Box 22, New York, NY 10065. This work was presented as a poster at the American Society for Radiation Oncology Meeting, October 2025.

## Abstract

Sentinel lymph node biopsy (SLNB) is increasingly omitted in early-stage breast cancer, often prompting whole-breast irradiation (WBI). We evaluated partial-breast irradiation (PBI) without axillary surgery among 78 clinically node-negative patients (median age 75) treated from 2014–2022. After 53-month median follow-up, no ipsilateral, regional, or distant recurrences occurred. These results demonstrate excellent outcomes and suggest PBI is a feasible, safe alternative to WBI when SLNB is omitted.

## Introduction

Significant efforts have been made in recent years to reduce the morbidity of regional nodal management among patients with early-stage breast cancer. Approximately three decades ago, technical advances enabled sentinel lymph node biopsy (SLNB) to replace the long-held standard of axillary lymph node dissection (ALND).^1^ In contemporary practice, even SLNB has been omitted for older patients with early-stage disease, based upon exceedingly low rates of axillary involvement.^2-4^

More recently, landmark studies have expanded on these efforts, eliminating axillary surgery entirely. The SOUND (Sentinel Node vs Observation After Axillary Ultra-Sound) and INSEMA trials evaluated omission of SLNB in a broad group of patients with early-stage breast cancer, demonstrating favorable oncologic outcomes.^5,6^ This paradigm shift in surgical practice has generated uncertainty with regard to the implications for adjuvant radiotherapy (RT). Most patients on SOUND and INSEMA received whole beast irradiation (WBI), although they would have been eligible for partial breast irradiation (PBI) in the setting of a negative SLNB. Similarly, the landmark trials demonstrating the safety of PBI largely mandated SLNB or ALND, such that PBI outcomes are largely undefined in the absence of surgical nodal evaluation.^7,8^

Here, we report oncologic outcomes among patients who received PBI despite undergoing lumpectomy without SLNB or ALND.

The study cohort comprised 78 patients (median age: 75 years; range 50-91), with significant representation below 70 years of age (Table 1). Most patients had invasive ductal carcinoma (79%), with only 8 (10%) cases of invasive lobular carcinoma. Nearly all patients had T1 disease (N=67, 86%), with few cases of T1mic (N=10; 13%) or T2 disease (N = 11; 14%). All patients were clinically node negative by physical examination. Most had low or intermediate grade disease (N=59, 76%), with only 11 (14%) exhibiting high grade disease. Most of the cohort had estrogen-receptor positive (ER+) (N=72, 92%), HER2 negative disease (N=71, 91%) and received endocrine therapy (77%). Only one patient underwent pre-operative axillary ultrasound.

**Table 1:**
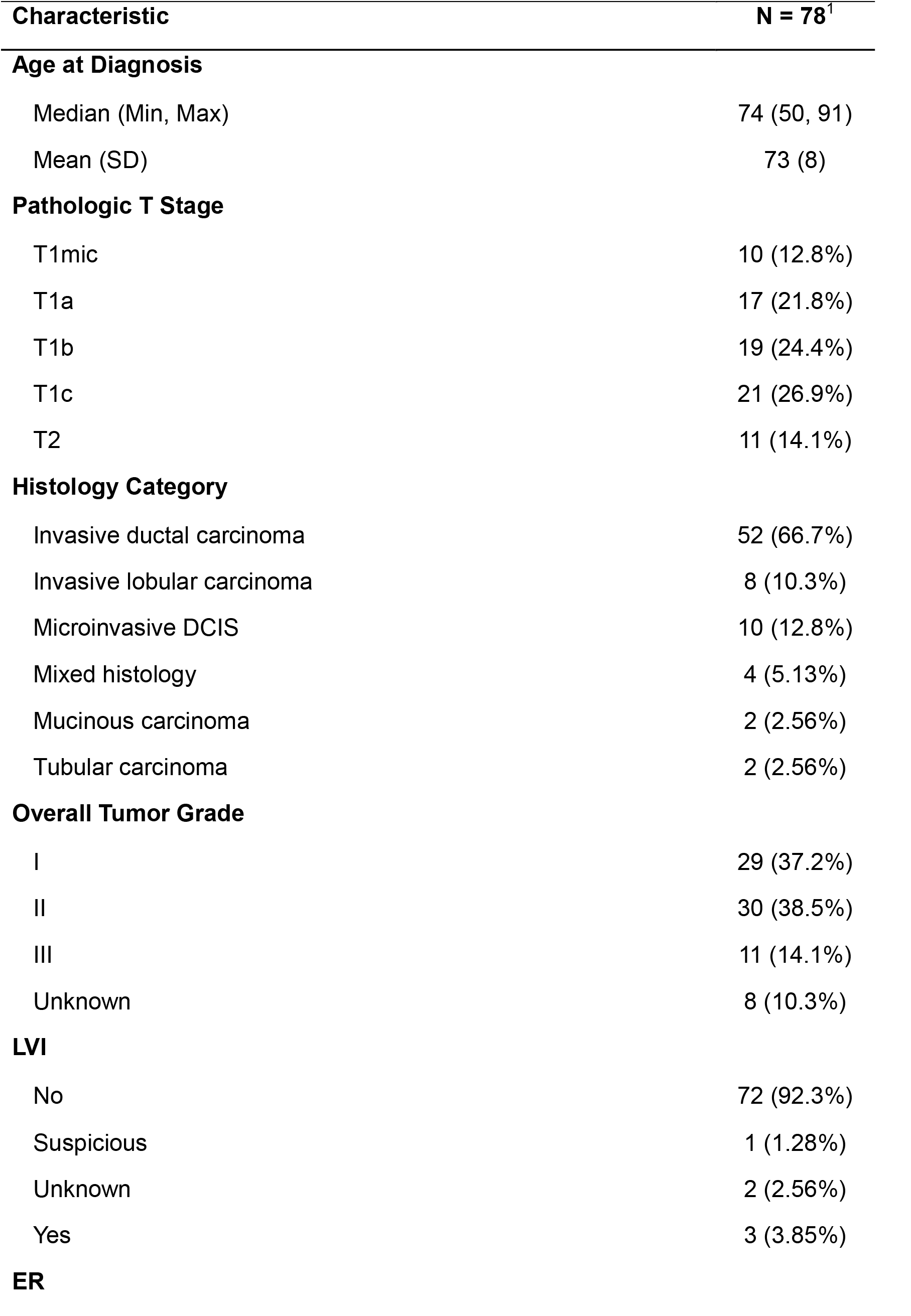
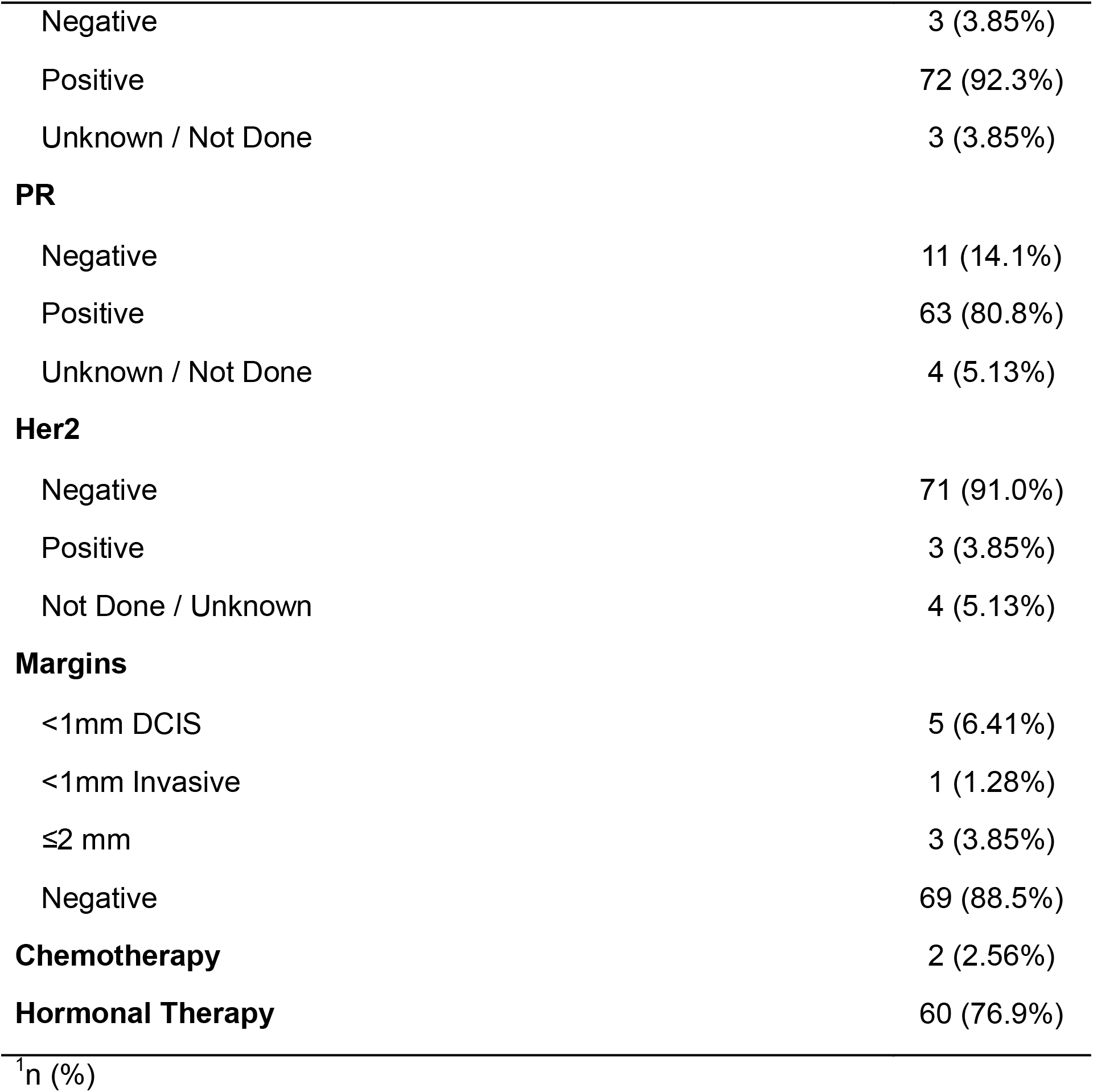
Patient characteristics.

The majority of patients (N=47, 60%) did not meet criteria for omission of adjuvant RT per CALGB 9343 criteria, most commonly due to not receiving endocrine therapy. 18 patients had ER+ disease but declined endocrine therapy, or had ER-disease, 14 patients were younger than 70, 11 had T2 disease, 7 had other cancer diagnoses within the 5 years preceding their breast cancer diagnosis, and 6 had synchronous contralateral breast disease.

At a median follow-up of 53 months, no local, regional or distant invasive or *in situ* recurrences were observed (Figure 1). Two patients experienced a contralateral breast cancer, yielding a 4-year cumulative incidence rate of 1.3% (95% confidence interval 0.1-6.3). Four non-breast-cancer-related deaths were noted during the follow-up period, at 4.2, 5.5, 6.4 and 7.1 years from diagnosis.

**Figure 1:**
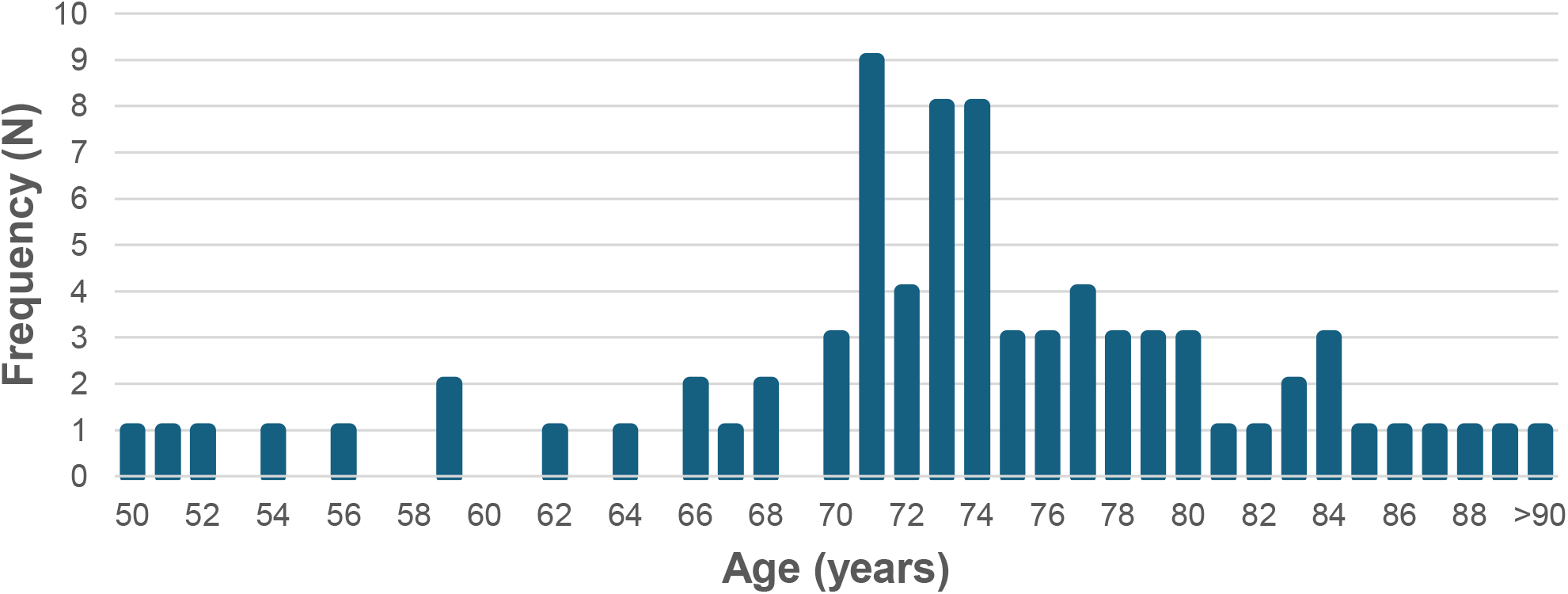
Cohort distribution of age at diagnosis

**Figure 2:**
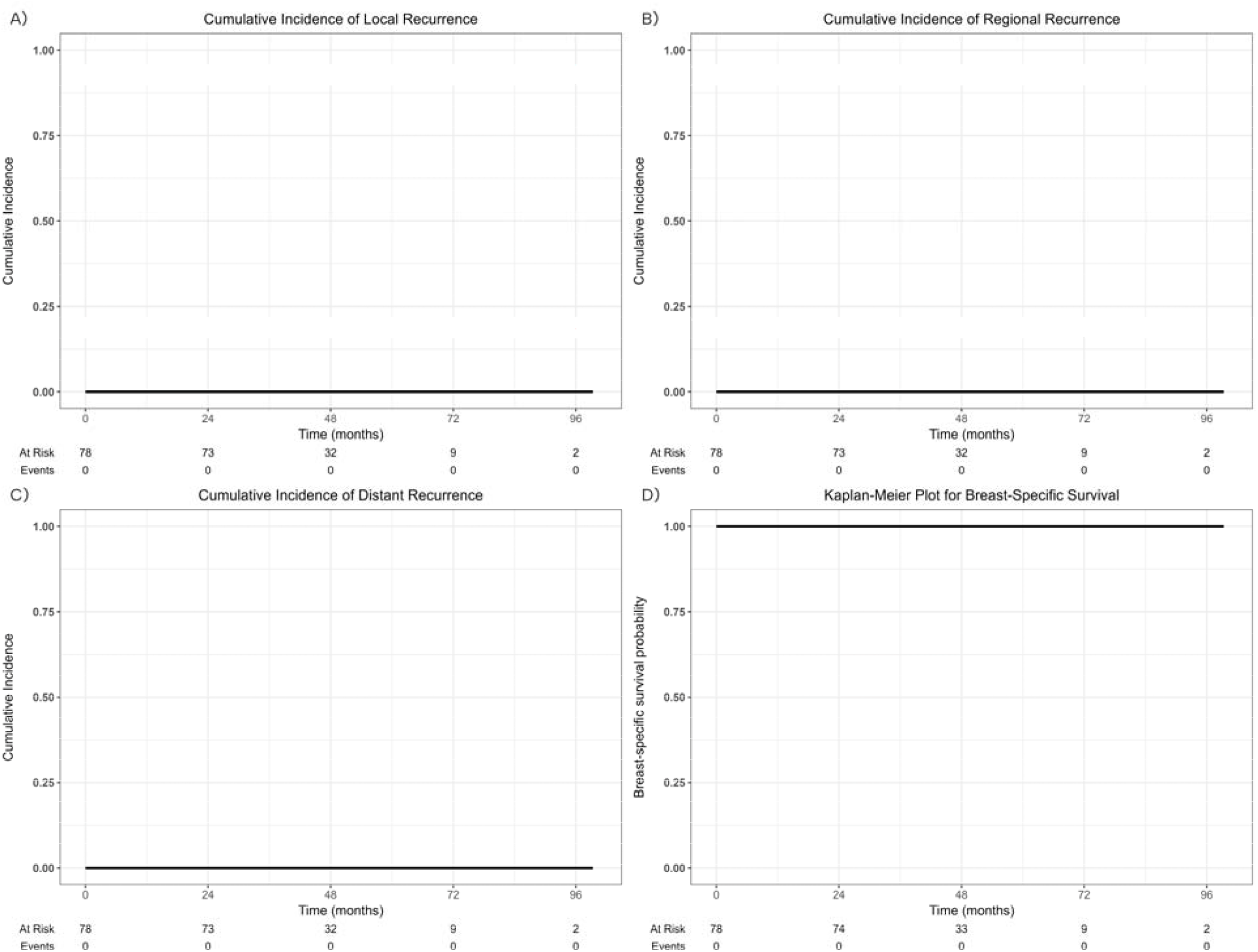
Oncologic outcomes, including cumulative incidence of A) local recurrence B) regional recurrence C) distant recurrence, and D) breast cancer-specific survival.

Here, we demonstrate excellent oncologic outcomes among patients with clinically node-negative, early-stage breast cancer managed without surgical axillary nodal evaluation and with PBI. At a median follow-up of 40 months, we observed no local, regional or distant recurrences, nor any breast cancer-related mortality. These findings suggest that PBI may remain a feasible treatment option, even in the absence surgical evaluation of the axilla. It is noteworthy that while this cohort had overall low risk breast cancer, these were largely patients for whom adjuvant RT was warranted, as the majority did not meet CALGB 9343 criteria for omission.

Axillary management in early breast cancer has progressively moved toward less invasive approaches. Historically, ALND was standard for all patients, despite its associated morbidity, particularly lymphedema. SLNB subsequently emerged as a less invasive alternative with comparable oncologic outcomes, representing a significant de-escalation in surgical management.^9-11^ Current guidelines from the American Society of Clinical Oncology, National Comprehensive Cancer Network, and Society of Surgical Oncology have begun to permit omission of even SLNB for low-risk patients—typically those ≥70 years old with T1N0, ER+, HER2-negative breast cancer.^2-4^

Recently, two landmark trials, SOUND and INSEMA, further questioned whether surgical staging of the axilla is necessary for early-stage disease.^5,6^ SOUND was a phase 3, randomized controlled trial of 1463 cN0, pT1 patients managed with pre-operative axillary ultrasound to demonstrate cN0, and lumpectomy. Patients were randomized to standard of care SLNB, versus no axillary surgery, ultimately demonstrating non-inferiority of omission of SLNB with regard to distant disease-free survival, disease-free survival, and OS.^5^ INSEMA, a similar randomized non-inferiority trial also evaluated omission of SLNB. 5502 eligible patients with cN0, T1-2 breast cancer were treated with lumpectomy with preoperative axillary ultrasound.

Omission of SLNB was non-inferior to SLNB with regard to invasive disease-free survival and, was associated with decreased lymphedema, improved arm mobility, and decreased pain with arm and shoulder movement.^6^

While these trials support surgical de-escalation, they predominantly used WBI in the adjuvant setting - INSEMA mandated WBI, and approximately 90% of patients on SOUND received it.^5,6^ For many patients, this represents a potential escalation of RT compared to PBI which would otherwise offer several advantages including smaller irradiated volumes, favorable toxicity profiles, potentially improved cosmesis, and reduced treatment burden.^12^ INSEMA included a preplanned evaluation of axillary RT dose, and found that at least half of analyzed patients received ≥80% of the prescribed RT dose to level 1 of the axilla, although dose to the axilla did not differ between the arms of the trial.^13^ There is hypothetical concern that in the absence of SLNB, WBI is addressing unresected microscopic disease in the low axilla. No data are available regarding outcomes of the subgroup of SOUND patients managed with PBI (in the form of intraoperative RT), rather than WBI, and to our knowledge, there is no literature regarding use of PBI in the absence of axillary nodal evaluation. Our study represents the first assessment of PBI in this setting, and suggests it may be a promising approach, worthy of further investigation. This work contributes to a growing body of literature suggesting that eligibility for PBI may be safely expanded in the future.^14-16^

Our findings must be interpreted in the context of the study design. The cohort was modestly sized and predominantly comprised older patients for whom physicians were appropriately less concerned about the omission of SLNB. The retrospective nature of our study is necessarily susceptible to bias and confounding, as omission of SLNB was at the discretion of the treating surgeon, and use of PBI was at the discretion of the treating radiation oncologist.

This resulted in a low-risk breast cancer cohort. However, as noted above, we did include a subset of patients younger than 70, and most patients did meet criteria for addition of adjuvant RT. Given the nature of our study population, our conclusions should be generalized to younger or higher-risk patients with caution. This cohort was composed predominantly of patients with ER+ breast cancer, a diagnosis with a long natural history and documented recurrences many years after initial diagnosis. Therefore, a limitation of this study is the relatively short follow up period of just over 4 years. Further follow-up among larger cohorts will be necessary to validate the excellent rates of disease control observed here.

Among patients with early-stage, clinically node negative breast cancer managed with breast conservation without SLNB, PBI was associated with excellent oncologic outcomes, as no recurrences were observed. Thus, following the contemporary de-escalation of axillary surgery, this population may remain eligible for PBI. The omission of surgical nodal evaluation need not mandate an escalation in locoregional radiotherapeutic approach.

## Methods

We identified all breast cancer patients treated at our center from 2014 to 2022 with lumpectomy and PBI, but who did not undergo SLNB or ALND. Salient clinicopathologic and treatment parameters were collected from the medical record. Patients were further classified by their suitability for RT omission.

The typical PBI regimen has been previously described.^17^ In brief, 40Gy was delivered in 10 daily fractions using external beam RT. The surgical cavity was delineated on a CT-simulation scan, with the aid of surgical clips placed intraoperatively. A clinical target volume (CTV) was generated as an isotropic expansion of 1.5-2 cm on the surgical cavity, limited anteriorly to within 0.5 cm below the skin surface and posteriorly to the anterior surface of the chest wall/ribs, followed by a 0.5 cm isotropic expansion to yield the planning treatment volume (PTV). Daily image guidance with kV imaging was employed, matched on surgical clips.

Primary endpoints included rates of ipsilateral breast tumor recurrence (IBTR), contralateral breast recurrence, regional recurrence (RR), or distant recurrence (DR), as well as overall survival (OS). Follow-up was defined as the time from diagnosis to the first recurrence, death, or last follow-up. Cumulative incidences of each event type were estimated with other events and death as competing risks. OS was estimated using the Kaplan Meier method. This study was approved by the Memorial Sloan Kettering Cancer Center institutional review board. The requirement for written consent was waived in light of the retrospective and de-identified nature of this study.

## Data Availability

All data produced in the present study are available upon reasonable request to the authors

## (1) Data Availability

The data that support the findings of this study are available from the corresponding author upon reasonable request.

## (2) Code Availability

(where applicable); N/A

## (3) Acknowledgements

(note: statement should declare any Funding for the work); **Funding/Support:** This study was supported in part by the Achar Meyohas Family, the Lois Green Fund, the Rose-Margulies Family Research Fund and NIH/NCI Cancer Center Support Grant No. P30CA008748.

## (4) Author Contributions

(individual contributions denoted by each author’s initials);All authors contributed to the conception and writing of this manuscript

